# GenPK Suite: An Integrated Digital Platform for Phenotypic Data Collection, 3D Facial Imaging, and Research Workflow Management in Rare Diseases

**DOI:** 10.1101/2025.11.18.25340411

**Authors:** Sikandar Farooq Akbar, Abdur Rashid, Ijaz Anwar, Alexandre Reymond, Federico Santoni, Muhammad Ansar

## Abstract

Accurate and secure collection of genetic samples and associated phenotypic data is essential for advancing rare disease research, yet existing workflows often remain fragmented across paper records, electronic questionnaires, and laboratory information systems. To address these limitations, we developed the GenPK Suite, an offline-capable digital platform that integrates family-wise recruitment, disorder-specific questionnaires, digital consent, pedigree capture, barcoded biospecimen tracking, and three-dimensional (3D) facial imaging. The system was deployed in field settings in Pakistan.

During pilot implementation, the platform enabled the recruitment of 121 families encompassing more than 150 individuals, resulting in 150 barcoded biospecimens and 50 high-resolution 3D craniofacial scans. Data completeness across mandatory fields exceeded 90%, while offline-to-cloud synchronization succeeded in >95% of encounters within 24 hours. Laboratory accession confirmed end-to-end traceability, with DNA quality and quantity metrics returned for samples. User feedback highlighted reduced paperwork burden and greater procedural consistency, with staff reporting fewer transcription mistakes and fewer manual linkage steps compared with their previous paper-based workflow.

By embedding technical safeguards aligned with ISO/IEC 27001 and 27701 and enforcing role-based access, the GenPK Suite demonstrated secure and practical feasibility for international rare disease research. These results show that integrated digital infrastructures can enable scalable recruitment and phenotyping across both high-resource and remote field environments.

## Introduction

Rare diseases are estimated to affect between 6% and 8% of the world’s population, an astonishing figure that translates to more than 300 million individuals worldwide (Boycott et al., 2017). Although rare conditions collectively impose a substantial burden, the pace of discovery has not matched that seen for common disorders. A persistent bottleneck lies in assembling cohorts that are both adequately powered and deeply phenotyped so that genetic and clinical inferences are credible. This demands both secure custody of biospecimens, carefully curated pedigrees, longitudinal histories, and structured clinical metadata (Posey et al., 2019). In practice, however, many groups still rely on paper questionnaires or a patchwork of partially compatible digital systems. This often leads to human errors, inefficiencies in storage, difficulties in standardization, and poor interoperability between clinical records, phenotypic assessments, and laboratory pipelines (Wilkinson et al., 2016). Our own experience supported by recent genomic and informatics studies confirms that even when data collection is rigorous, the lack of integration across systems makes harmonization extremely time-consuming and increases the risk of transcription or linkage discrepancies (Mattioli et al., 2025; Bassani et al., 2024; Zafar et al., 2025; Paracha et al., 2024; Mattioli et al., 2021).

Over the past decade, the adoption of digital infrastructures in biomedical research has accelerated, bringing with it important advances in data traceability, standardization, and security. Electronic data capture tools, laboratory information management systems (LIMS), and biobank management platforms have become increasingly common in both academic and clinical settings (Mascalzoni et al., 2015). These systems have unquestionably improved many aspects of research workflows. Yet they tend to be designed for one stage of the process rather than for the continuum of clinical and laboratory activities. REDCap (https://project-redcap.org/), for instance, is highly valued for questionnaire-based data entry and has the advantage of being customizable and relatively easy to deploy, but it does not natively integrate with barcode-based sample tracking or three-dimensional phenotyping. By contrast, laboratory information management systems (LIMS) excel inside the lab-accessioning, inventory control, aliquoting, and process tracking, but they rarely reach upstream to recruitment, e-consent, or pedigree capture. The consequence is predictable: teams juggle parallel tools, reconcile records by hand, and accept residual mismatches across datasets. These discrepancies can become more prominent in cross-border collaborations and/or in low-resource environments, where limited budgets, uneven infrastructure, and variable staff expertise make it unrealistic to operate several specialized platforms in tandem.

Another dimension that remains comparatively underused is advanced phenotyping. Three-dimensional (3D) facial imaging has shown strong promise for syndromes with craniofacial involvement, capturing subtle morphological patterns that aid clinical interpretation and can surface otherwise hidden genotype–phenotype relationships (Hammond et al., 2020; Pantel et al., 2020). The approach has proven value in specialist centers; nevertheless, broader diffusion has been hampered by equipment costs, training requirements, and the absence of native links to recruitment, consent, or sample workflows. Field observations mirror this: 3D scans can be acquired successfully, yet without direct integration into intake and laboratory pipelines, throughput stalls and scaling becomes difficult. In effect, the method remains confined to small, expert-led studies rather than the multi-institutional consortia needed for discovery at scale.

The same issues are magnified in remote or low-resource settings, where much of the rare-disease burden resides. Connectivity may be intermittent or absent, making cloud-only systems impractical. Data entry is then deferred until a connection is available or recorded in ad hoc spreadsheets and paper logs. Predictably, fragmentation and loss follow, and underrepresented communities are edged out of genomic cohorts. This under-participation, documented in international collaborations that aim for diversity (Mascalzoni et al., 2015), creates a feedback loop: populations with the greatest unmet needs remain sparsely represented, limiting the discovery of variants most relevant to their care.

Taken together, these realities argue for a unified, modular, and secure digital infrastructure that spans recruitment, phenotyping, and laboratory analysis. The GenPK Suite was developed with this goal in mind through collaboration among Jules-Gonin Eye Hospital, the University of Lausanne, and ICASTA LLC (International Center for AI, Software, and Emerging Technologies of America). Designed for field campaigns, the platform runs on Amazon Web Services (AWS) to provide elastic scaling, robust security controls, and tightly governed access across sites. Its architecture combines three complementary components: (i) a mobile application for structured intake, consent, and metadata capture; (ii) an iOS application for 3D craniofacial imaging; and (iii) a role-based web portal that manages users, tracks samples, and links laboratory workflows.

This paper details the design, implementation, and deployment of the GenPK Suite across distinct operational contexts. The account is intentionally practical: it shows how an integrated, offline-capable, and compliance-aware platform can make rare-disease research more secure and efficient, while remaining scalable across institutions. Beyond facilitating data capture, the Suite offers an example of how clinical, laboratory, and phenotypic domains can be woven into a single working ecosystem to support durable, global collaboration.

## Methods

### Evaluation Design

To assess the feasibility and performance of the GenPK Suite under routine operating conditions, pilot deployments were conducted across both high-resource and low-resource contexts. The system was implemented in field settings in Pakistan, where connectivity and device availability are more constrained.

Participants and data volume. A total of 121 families comprising more than 150 individuals were recruited using the GenPK Collect mobile application during its pilot phase. This process generated approximately 150 barcoded biospecimens and 50 three-dimensional (3D) craniofacial scans, each linked to unique individual identifiers and corresponding consent records.

**Primary endpoints.** The main measures of feasibility were:

- **Data completeness**: proportion of encounters in which all mandatory fields were captured.
- **Questionnaire coverage**: percentage of disorder-specific questionnaires ≥95% complete.
- **Offline synchronization success**: proportion of records successfully uploaded to the central server within 24 hours of capture.
- **Barcode linkage integrity**: rate of duplicate identifiers or mis-linked samples per 1,000 barcodes.
- **System stability**: proportion of crash-free collection sessions.
- **3D scan adequacy**: percentage of scans rated as sufficient for downstream morphometric analysis.
- **Laboratory accession turnaround**: median time from collection to laboratory confirmation of receipt.

### Secondary endpoints

Additional measures included the proportion of samples with DNA quality and quantity metrics recorded, the completeness of audit logs (≥3 timestamped events per sample), and qualitative feedback on usability from collectors, clinicians, and laboratory staff.

### Analysis

Outcomes were summarized descriptively using counts, percentages, and medians with interquartile ranges (IQRs). Results were stratified by deployment context (hospital vs. field) to evaluate generalizability across environments.

### Standards Compliance

The GenPK Suite was developed in alignment with internationally recognized information security and privacy management standards. Technical and organizational safeguards were mapped to specific ISO/IEC controls to ensure secure handling of clinical and genetic data.

- **Information security management.** Core security practices were aligned with **ISO/IEC 27001** and **ISO/IEC 27002**, including risk assessment, access control, cryptographic key management, and incident response planning.
- **Cloud and data governance.** Cloud deployment was implemented in accordance with **ISO/IEC 27017** (cloud-specific security controls) and **ISO/IEC 27018** (protection of personally identifiable information in public cloud environments).
- **Privacy management.** Data minimization, subject access rights, and retention policies were guided by **ISO/IEC 27701**, extending privacy information management (PIMS) requirements to the research context.
- **Access control.** Role-based access control (RBAC) adhered to ISO/IEC 27001 Annex A.9, ensuring that users accessed only the minimum information required for their roles (e.g., laboratory staff restricted to sample identifiers).
- **Data protection in transit and at rest.** All communications were encrypted using TLS, while structured and unstructured data were stored under AES-256 encryption with keys managed through AWS Key Management Service (KMS), consistent with ISO/IEC 27001 Annex A.10.
- **Audit and traceability.** Logging and monitoring followed ISO/IEC 27001 Annex A.12, ensuring that every critical event, such as consent capture, barcode assignment, and laboratory accession—was timestamped, authenticated, and preserved in immutable form.

By embedding these controls, the GenPK Suite was designed to align closely with international information security and privacy management standards. It should be noted, however, that this alignment reflects internal architectural and procedural mapping to ISO/IEC 27001, 27017, 27018, and 27701; formal third-party certification and independent penetration testing had not yet been performed at the time of this pilot deployment. A summary of system elements and their corresponding ISO/IEC controls is presented in Table 1.

**Table 1:** Summary of system elements and their corresponding ISO/IEC controls.

| System element | ISO/IEC control family | Implementation in GenPK Suite |
| --- | --- | --- |
| Data encryption in transit | 27001 A.10, 27002 §10 | TLS enforced for all app–server traffic |
| Data encryption at rest | 27001 A.10, 27018 | AES-256 with AWS KMS key rotation |
| Access control | 27001 A.9 | RBAC with least-privilege hierarchy |
| Logging and audit | 27001 A.12 | Immutable, time-synced audit trails |
| Privacy management | 27701 | Data minimization, retention policies |
| Cloud security | 27017, 27018 | VPC isolation, GuardDuty monitoring |

### System Architecture

The GenPK Suite was implemented as a modular digital ecosystem designed to support rare disease research across both hospital and field environments. The platform comprises three integrated components: GenPK Collect, a mobile application for structured intake and sample capture; GenPK Scan, an iOS-based application for three-dimensional (3D) craniofacial imaging; and a secure web portal for role-based administration, laboratory accessioning, and data oversight (Figure 1).

**Figure 1.**
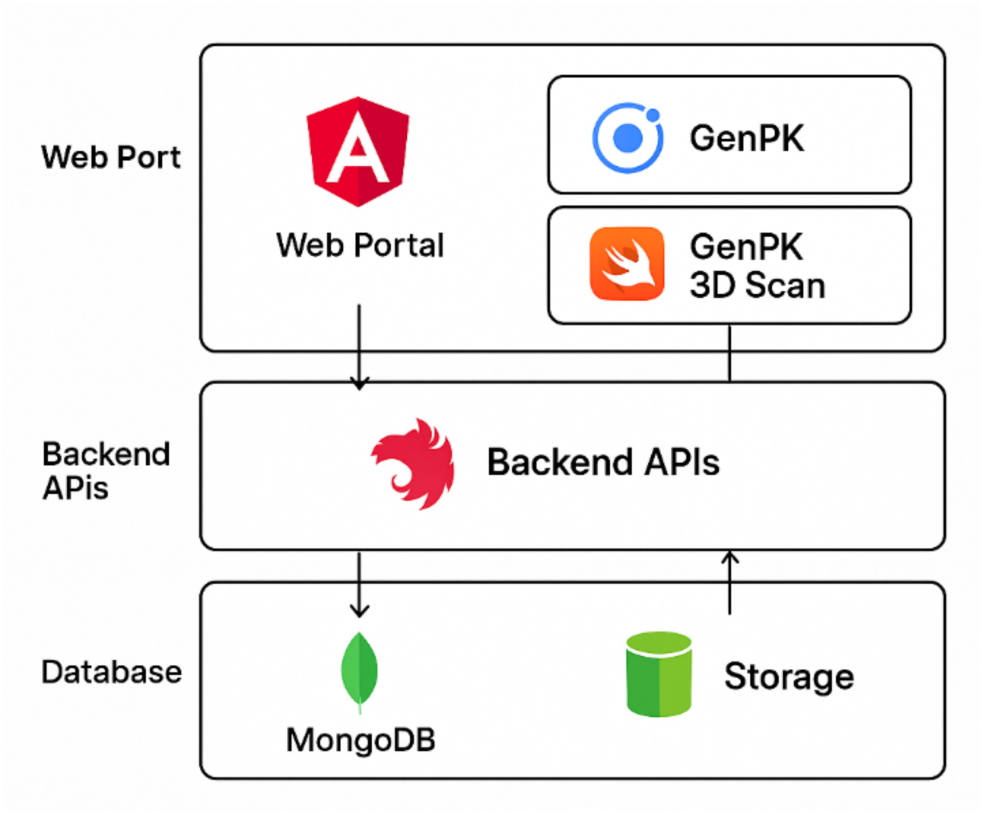
System architecture overview of the GenPK Suite, illustrating the three major components (GenPK Collect, GenPK Scan, and the Web Portal) connected through backend services and a centralized database.

All components interface through a centralized backend that supports offline-first operation. Data collected in the field are encrypted and stored locally, then synchronized to the central server when connectivity is restored. This model ensures continuity of recruitment in low-resource contexts while maintaining traceability across identifiers, consent forms, barcoded biospecimens, and 3D scans.

To enable multi-institutional use, the platform incorporates a multi-tenant structure in which administrators can register institutions, assign roles, and manage data access hierarchies. Each entity, families, individuals, samples, scans, is linked to a unique identifier, providing interoperability between phenotypic, clinical, and laboratory workflows within a unified environment.

Figure 1 provides an overview of the GenPK Suite architecture, showing the three major components (Collect, Scan, Portal) and their integration through backend services and database layers. Figure 2 depicts the detailed data flow and security model, highlighting offline capture, encrypted synchronization, role-based access, and alignment with cloud-native safeguards. Together, these schematics illustrate how the system bridges intake, phenotyping, and laboratory workflows within a secure and offline-capable infrastructure.

**Figure 2.**
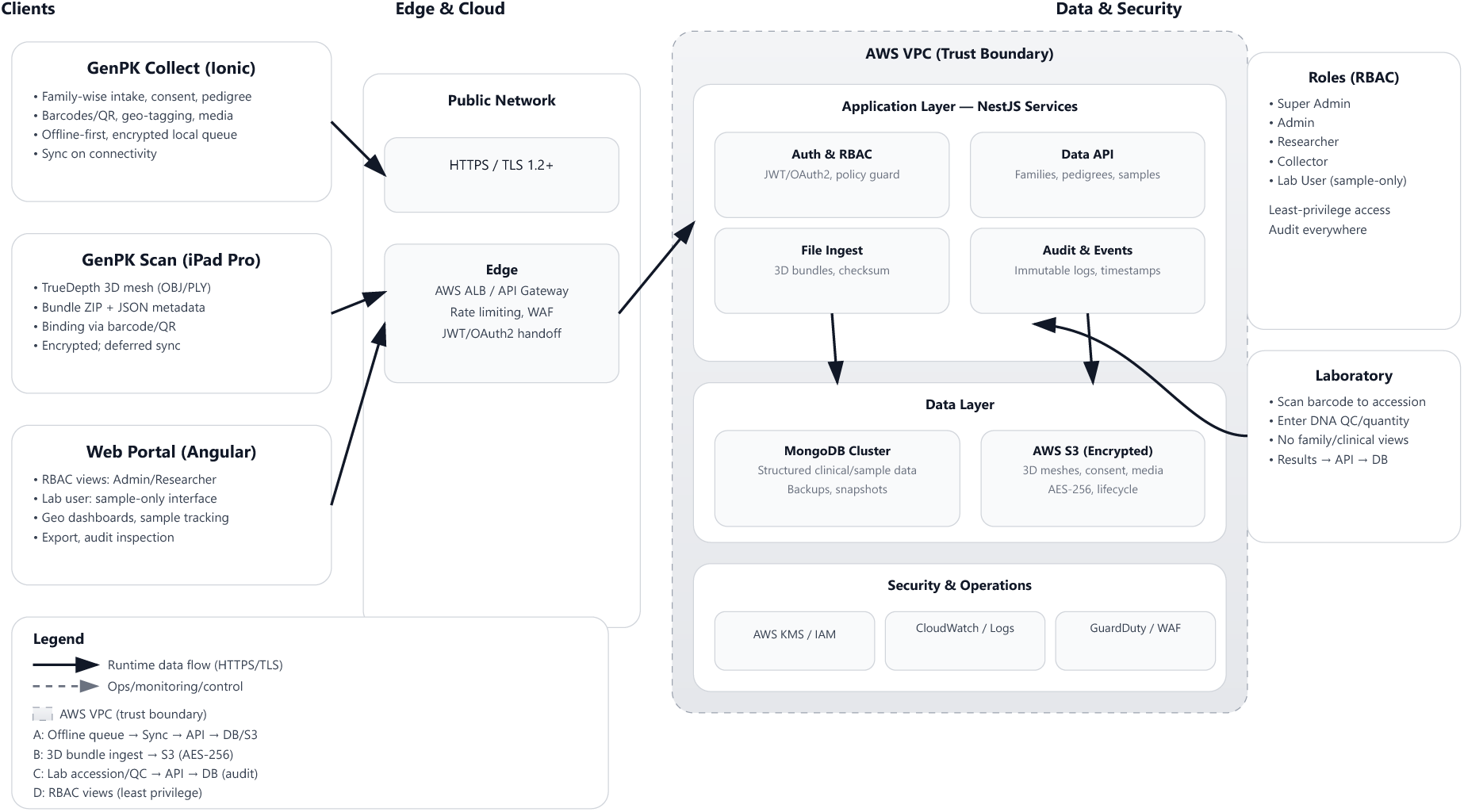
Detailed system workflow showing client applications, backend services, data storage, and security operations. The diagram emphasizes offline-first data capture, encrypted synchronization, role-based access control, and compliance with cloud security standards.

The data flow between components follows a hub-and-spoke model, in which mobile and scanning applications operate as decentralized collection tools, capable of functioning offline, and later synchronize with the central server when connectivity is restored. This architecture ensures resilience in remote settings where internet access is intermittent, while still maintaining consistency and traceability once synchronization occurs. Each data entity, such as family identifiers, individual records, questionnaires, consent forms, barcodes, and 3D models, is linked via unique identifiers, ensuring interoperability between different modules of the system.

To support collaboration at scale, the system incorporates a multi-tenant structure. This allows administrators to onboard institutions, assign roles (e.g., Super Admin, Admin, Researcher, Collector, Laboratory), and restrict access to data based on predefined hierarchies. Integration between the 3D imaging module and the central data repository enables phenotypic data to be seamlessly linked with genetic and clinical records, thereby bridging previously fragmented workflows into a single cohesive infrastructure.

### GenPK Collect: Mobile Application

The GenPK Collect mobile application served as the primary interface for field workers to capture structured clinical and genetic data. Recruitment followed a hierarchical model: each family was assigned a unique identifier, and each individual within that family was linked through a nested identifier, allowing pedigrees, medical histories, and biospecimens to be systematically connected for downstream analyses.

The application supported disorder-specific questionnaires tailored to conditions such as intellectual disability, visual impairment, skeletal deformities, and neonatal metabolic disorders. When individuals presented with multiple phenotypes, the relevant questionnaires were dynamically loaded, ensuring comprehensive capture of overlapping features. Importantly, both questionnaires and informed consent forms are implemented as configurable templates that can be updated or replaced without modifying the core application. This allows new studies to import their own study-specific instruments, consent forms, or medical history modules, making the system adaptable for a wide range of research projects. Recruitment also included unaffected family members, providing internal controls for genotype–phenotype analyses.

Ethical compliance was ensured through an embedded digital consent module. Participants or guardians provided electronic consent directly on the device, and signatures were encrypted and permanently linked to the corresponding individual record. Each biological specimen (blood or saliva) was labeled with a barcode or QR code, cross-referenced to the individual identifier, thereby establishing end-to-end traceability from collection to laboratory accession.

Additional field-level utilities included automated recording of geo-coordinates, graphical pedigree construction, and secure capture of multimedia records such as photographs, videos, and scanned medical documents. These contextual data enriched phenotypic characterization and supported geographic diversity analyses while preserving confidentiality through role-based restrictions.

A central design principle was offline-first functionality. Data captured in remote settings were encrypted and stored locally on the device. Upon restoration of connectivity, records were automatically synchronized with the central server via secure channels. Modification of records required explicit approval: collectors could request temporary authorization from supervisors (Admin or Researcher), with changes logged and linked to time-limited access tokens. This mechanism balanced flexibility with data integrity and accountability. The recruitment and data capture workflow implemented in GenPK Collect is summarized in Figure 3.

**Figure 3.**
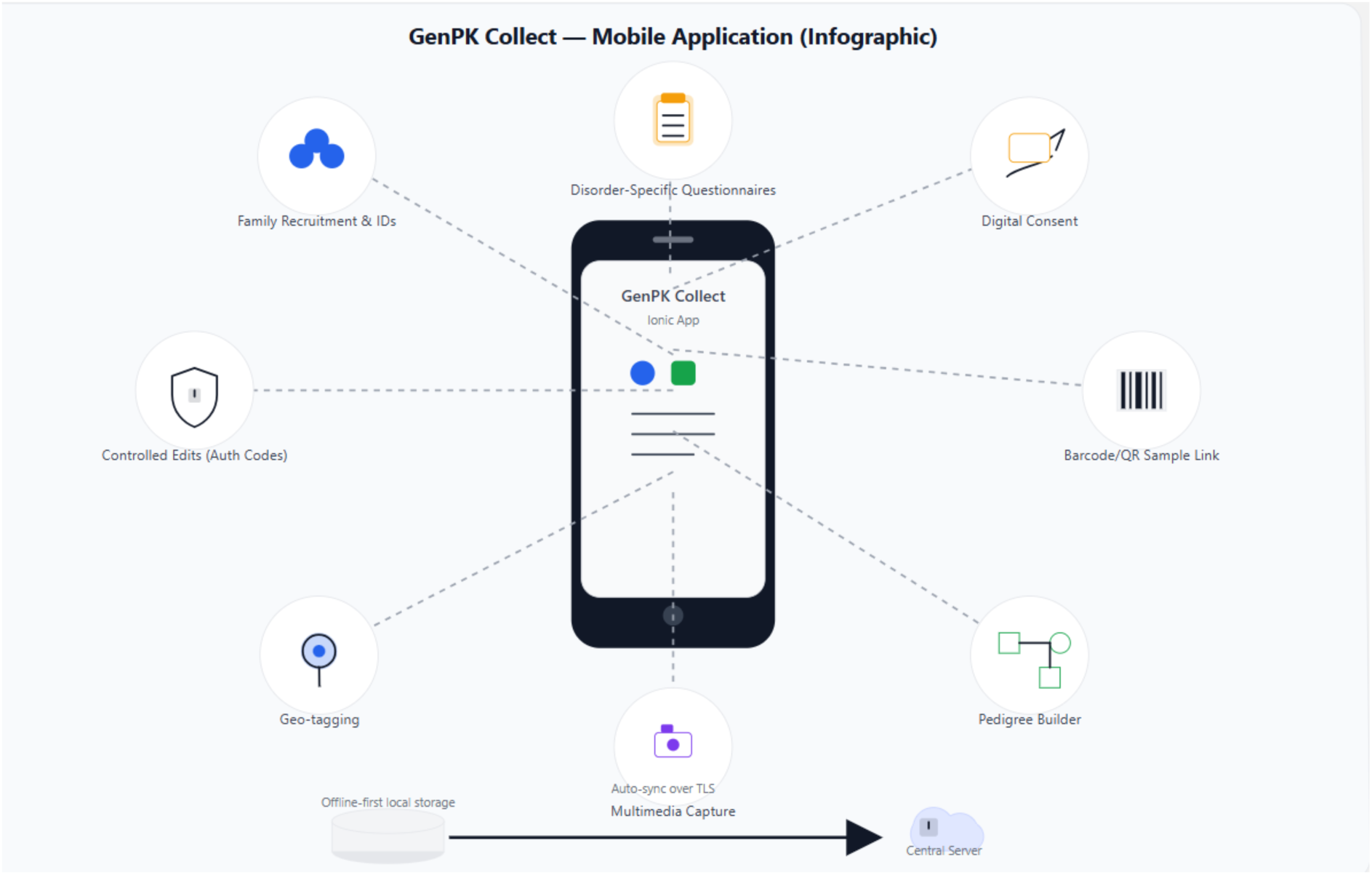
Workflow of the GenPK Collect mobile application, illustrating recruitment, digital consent, barcoded specimen capture, and offline synchronization for field-based deployment.

### Consent, Pedigree, and Geospatial Data Capture

Ethical compliance and contextual data capture were treated as core requirements in the design of the GenPK Suite. Three elements, digital consent, pedigree construction, and geospatial tagging, were integrated into the mobile and web applications to ensure that participant records were both ethically sound and analytically robust.

The digital consent module allowed participants or their guardians to review approved forms and provide electronic signatures directly on the device. Each consent record was encrypted at capture, time-stamped, and permanently linked to the individual identifier. These artifacts were retrievable through the audit trail, providing verifiable provenance during ethics review or inter-institutional data sharing.

Pedigree construction was implemented through an interactive tool in which families were assigned unique identifiers and individuals linked by structured relational fields. Graphical documentation of family relationships and health status ensured that affected and unaffected relatives could be linked to the same genetic metadata. The deliberate inclusion of unaffected family members created internal controls, thereby strengthening genotype–phenotype analyses and improving statistical interpretability.

Geospatial tagging recorded GPS coordinates at each collection event. To reduce the risk of re-identification, these data were stored in a logically separated partition of the database with access restricted to authorized researchers and administrators. The resulting geospatial dataset enabled visualization of recruitment diversity and provided a basis for geographic analyses of rare disease prevalence. The integration of consent, pedigree, and geospatial data into unified participant records is depicted in Figure 4.

**Figure 4.**
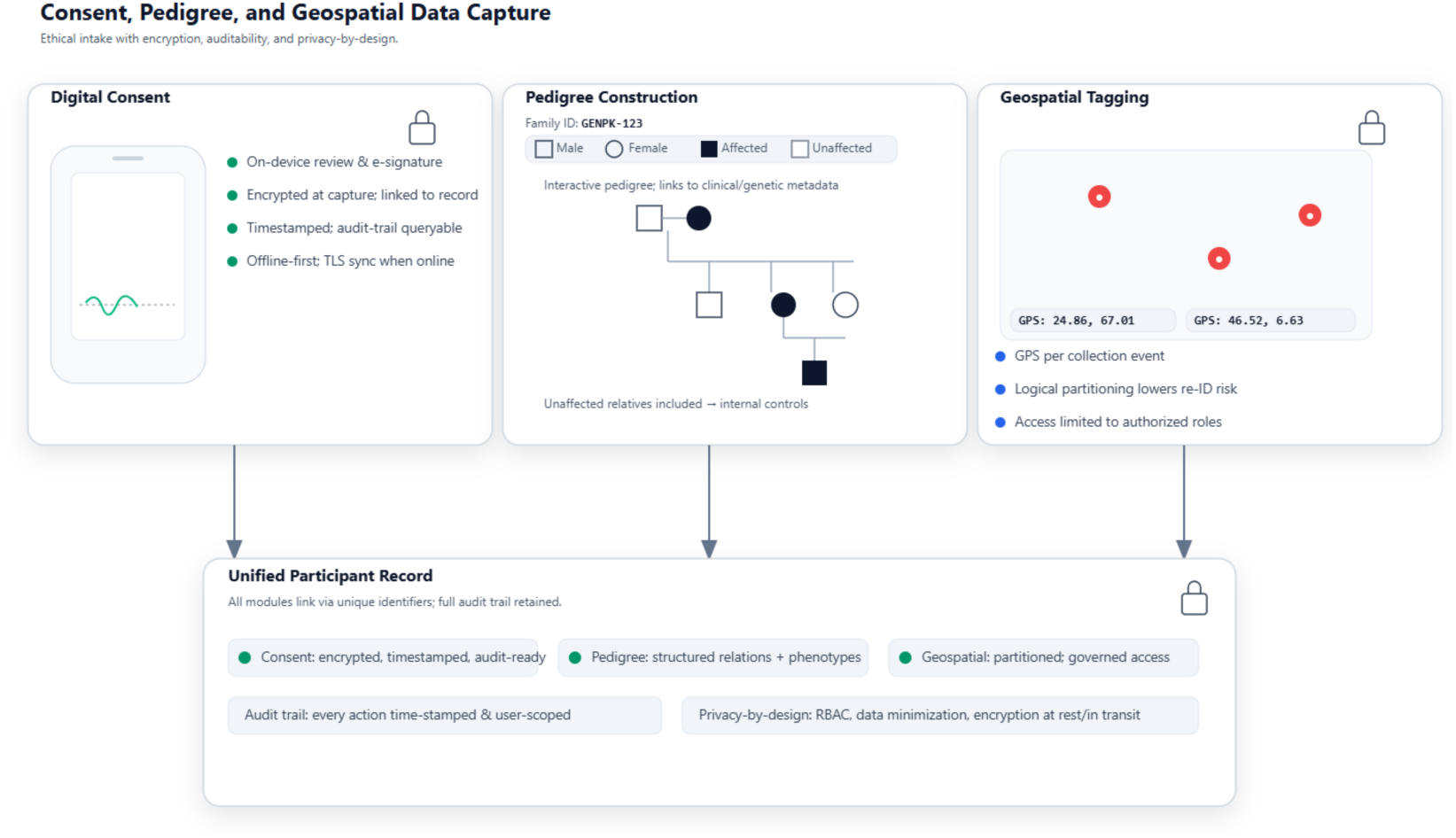
Consent, pedigree, and geospatial data capture in the GenPK Suite. The diagram illustrates how digital consent, interactive pedigree construction, and GPS tagging converge into unified participant records governed by role-based access and audit trails.

By combining these features, each participant record in the GenPK Suite linked consent, family context, and geospatial data under a unified identifier. This design enhanced participant protection, reproducibility, and the capacity to interrogate genetic variation within familial and geographic frameworks.

### GenPK Scan: 3D Imaging Application

The GenPK Scan module was developed as a companion iOS application to enable three-dimensional (3D) phenotyping within the broader GenPK Suite workflow. The application targets Apple iPad Pro hardware and leverages the built-in TrueDepth sensor to acquire dense, textured craniofacial meshes. The capture workflow is optimized for facial morphology but is region-agnostic and can be adapted for other anatomical regions (e.g., hands, feet, head circumference) when clinically relevant.

### Acquisition workflow

Each scan session begins with participant selection from the roster or by scanning the individual’s barcode/QR identifier, binding the capture directly to the correct digital record. Visual guidance is provided in real time using ARKit’s face tracking framework (Apple Inc.), which supplies pose and alignment cues, coverage indicators, and stability feedback. This reduced the need for rescans and shortened the training curve for new collectors in both clinical and field deployments.

### Data handling

Captured meshes are exported in standard 3D formats (OBJ, PLY) along with material descriptors and textures. Metadata describing device, pose, and capture time are stored in a JSON sidecar file. The files are packaged into ZIP bundles, encrypted locally using AES-256, and queued for deferred synchronization. Upon restoration of connectivity, background transfer occurs via HTTPS/TLS; file integrity is verified using checksums before assets are linked to the participant’s record in the central database.

### Integration

Unlike conventional workstation-based 3D imaging tools, GenPK Scan is natively embedded in the same environment used for recruitment, digital consent, and biospecimen barcoding. This tight coupling minimized hand-offs and prevented metadata drift between imaging and clinical data. In practice, this allowed phenotypic capture at the point of recruitment, in hospital clinics, rural health posts, or community outreach campaigns, without requiring a separate imaging pipeline. The complete workflow, from acquisition to secure synchronization and role-based access, is illustrated in Figure 5.

**Figure 5.**
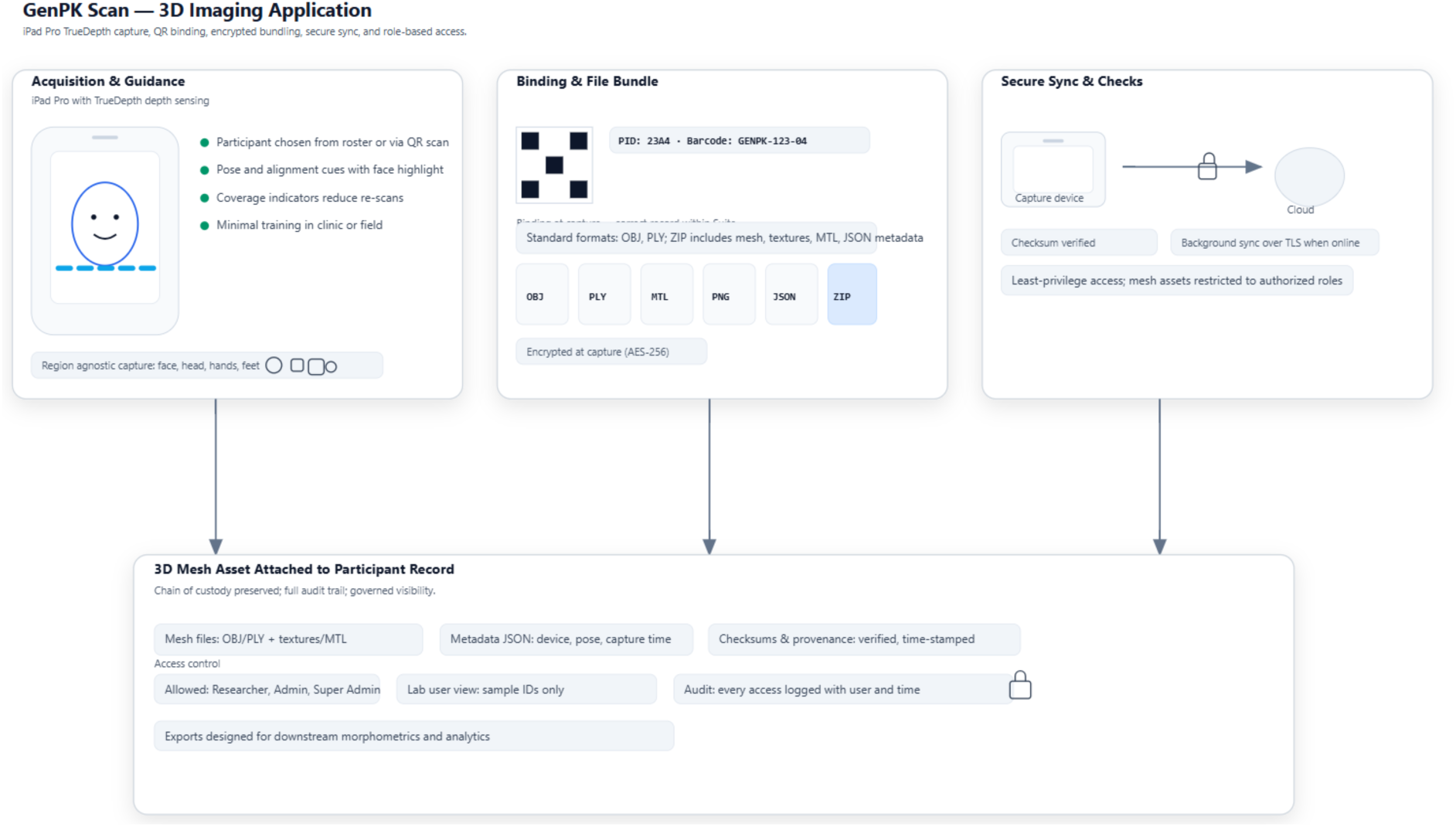
Workflow of the GenPK Scan module. The diagram illustrates acquisition with real-time guidance, binding of captures to participant identifiers, secure bundling of mesh and metadata, deferred synchronization via encrypted transfer, and integration with role-based access controls.

### Security and governance

Access to 3D meshes is governed by least-privilege, role-based controls (RBAC). Only Researcher, Admin, or Super Admin roles are permitted to view or download mesh assets. All access is logged with user ID and timestamp, ensuring auditability. Laboratory users remain restricted to sample identifiers only, without visibility of facial data or family records, in accordance with privacy-by-design principles.

### Web Portal and Role-Based Access System

The GenPK web portal served as the central hub for managing sample records, 3D imaging files, laboratory results, and user administration. The interface provided scalable data retrieval and seamless integration with the mobile applications, enabling harmonized oversight across hospital and field deployments.

A defining feature was the implementation of role-based access control (RBAC), which ensured data minimization and strict alignment of privileges with user responsibilities. Five hierarchical roles were defined:

- **Super Admin** – system-wide oversight, creation of Admin accounts, full visibility of data.
- **Admin** – management of researchers and collectors within their domain, creation of laboratory accounts.
- **Researcher** – supervision of designated collectors, with access to family records, pedigrees, and samples collected by their team.
- **Collector** – limited to families and samples personally recruited, including consent forms and barcoded specimens.
- **Laboratory User** – restricted to sample identifiers only, permitted to enter DNA quality and quantity metrics without visibility of family or clinical data.

In addition to access control, the portal incorporated geospatial dashboards that displayed families, laboratories, and collectors as map-based pins, supporting oversight of field campaigns and analyses of recruitment diversity. Operational modules included courier-style sample tracking with timestamped events, generation of barcode/QR labels, and a controlled modification request workflow in which collectors could obtain time-limited authorization to amend records with participant consent. Laboratory results were uploaded through the portal, version-controlled, and made available for secure download by researchers and administrators.

Through these combined features, the portal provided a secure, auditable chain of custody from sample collection to laboratory reporting. This workflow, together with the RBAC hierarchy and supporting modules, is illustrated in Figure 6.

**Figure 6.**
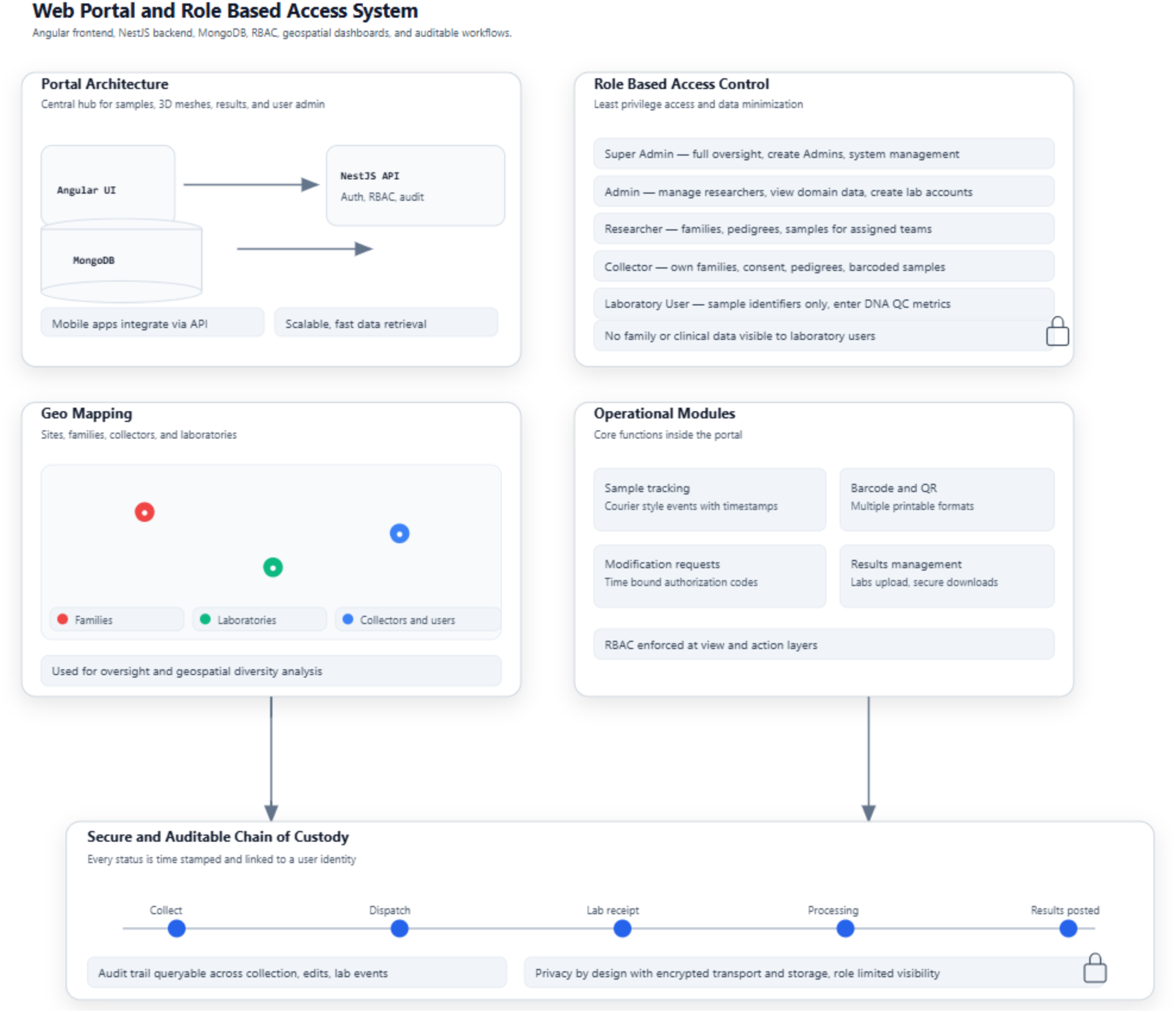
GenPK web portal and role-based access system. The schematic shows portal architecture, RBAC hierarchy, geospatial dashboards, operational modules, and the secure chain of custody from collection to laboratory results.

### Laboratory Workflow Integration

The GenPK Suite incorporated a laboratory workflow module to ensure complete traceability of biospecimens from collection through result reporting.

### Barcode binding at collection

At the time of collection, pre-printed barcode or QR labels generated by the GenPK portal were affixed directly to the blood or saliva tube by the collector. The label was then scanned with the GenPK Collect mobile application, which automatically linked the specimen to the individual’s record in the database. To prevent duplication or mislabeling, the application enforced single-use identifiers: once a barcode was bound to a participant, it could not be reassigned. A confirmation screen displayed the participant and sample IDs side-by-side, and the event was immediately written to the audit log with collector identity and timestamp.

### Dispatch and accession

Specimens were pre-assigned to registered laboratories through the mobile application or web portal prior to shipment. Upon receipt, laboratory personnel confirmed accession by scanning the barcode, with each accession event time-stamped and logged. Laboratory users operated under a sample-only view, restricted to identifiers and DNA quality metrics without access to family or clinical metadata, in line with data minimization principles.

### Processing and results integration

Laboratory staff entered DNA concentration and quality metrics directly into the portal. Entries were encrypted, version-controlled, and time-stamped, ensuring reproducibility and transparency. The modular design supports extension to additional assays, including whole exome sequencing (WES), whole genome sequencing (WGS), and Sanger sequencing. Results were linked to the sample identifier and made available for secure download by researchers and administrators.

### Tracking model

Sample tracking followed a courier-style model in which each transition—collection, dispatch, accession, processing, and result posting—was logged with user identity and precise timestamp. Researchers and administrators could visualize specimen progress through the portal, enabling oversight and rapid identification of delays or errors.

Together, these design features established a secure and auditable chain of custody consistent with international data protection standards. The complete laboratory workflow, from barcode binding to results integration, is illustrated in Figure 7.

**Figure 7.**
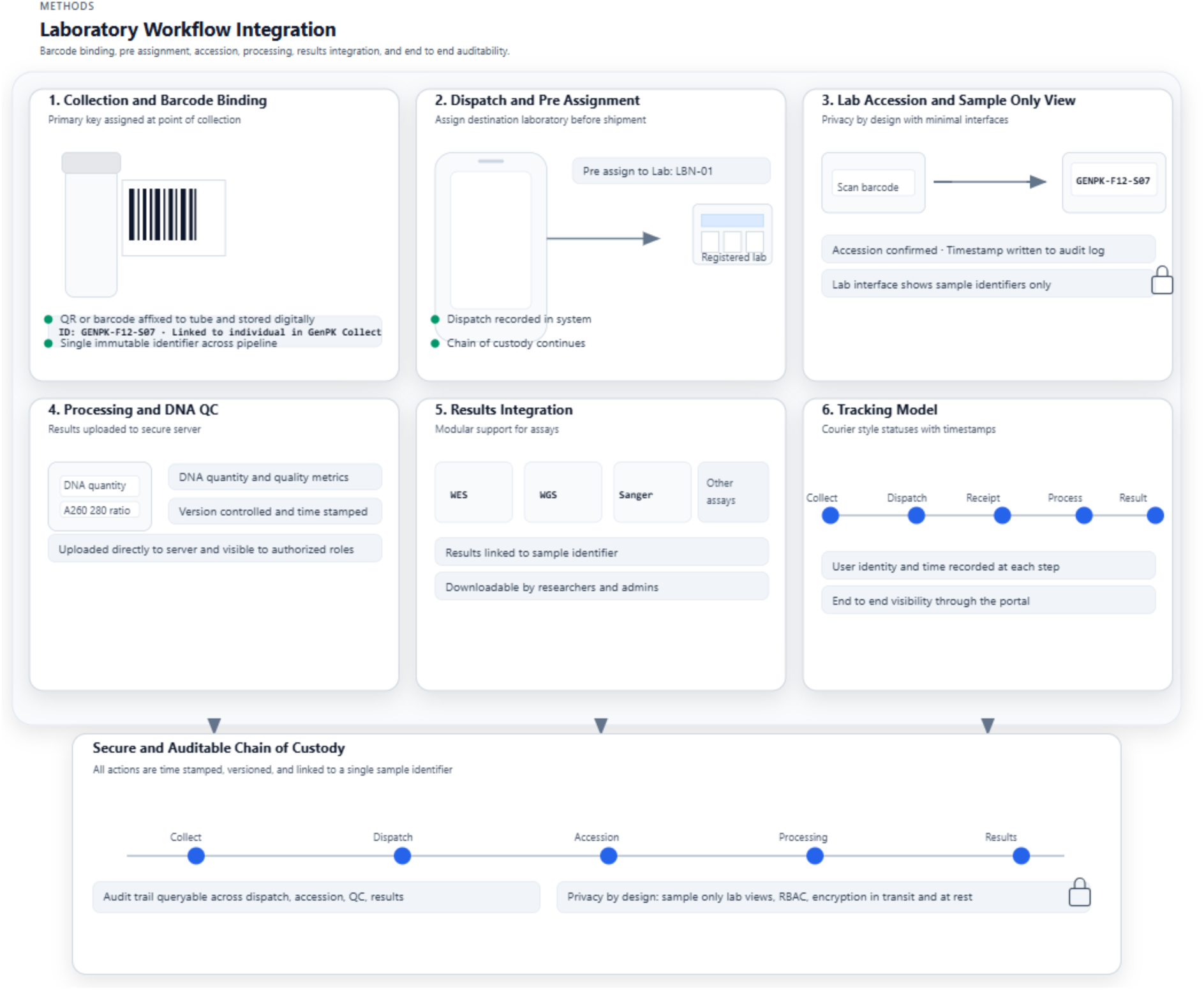
Laboratory workflow integration within the GenPK Suite. The schematic shows barcode assignment, laboratory pre-assignment, accession, DNA processing and quality control, results integration, and courier-style tracking with timestamps, forming a secure

### Data Security and Privacy Safeguards

Given the sensitivity of genetic and clinical information, data protection was embedded as a core design principle of the GenPK Suite. Security controls were mapped explicitly to international standards, including ISO/IEC 27001 (information security management), ISO/IEC 27002 (security controls), ISO/IEC 27017 (cloud security), ISO/IEC 27018 (PII in cloud), and ISO/IEC 27701 (privacy information management). The system incorporated layered safeguards spanning authentication, storage, privacy-by-design, auditability, and offline operation.

#### 1. Authentication and Authorization

User accounts were verified via email authentication before access was granted. Passwords were stored using bcrypt hashing with unique salts, consistent with ISO/IEC 27002 recommendations. Access followed role-based access control (RBAC) principles (ISO/IEC 27001 Annex A.9), ensuring that users (Super Admin, Admin, Researcher, Collector, Laboratory) interacted only with data required for their role.

#### 2. Data Transmission and Storage

All communication between mobile apps, web portals, and backend services was secured with TLS 1.2+ encryption (ISO/IEC 27001 A.10). Sensitive files, including consent forms and 3D scan bundles, were stored in AWS S3 with AES-256 encryption-at-rest, managed by AWS Key Management Service (KMS) with regular key rotation. Structured metadata were stored in MongoDB clusters configured with encryption-at-rest and automated backups. Disaster recovery and replication safeguards were aligned with ISO/IEC 27017 cloud practices.

#### 3. Privacy by Design

Access to personally identifiable information (PII) followed the principle of data minimization (ISO/IEC 27701). Laboratory users viewed only sample identifiers and quality metrics, while collectors were restricted to data they personally acquired. Any record modification required a time-limited authorization code issued by an Admin or Super Admin, accompanied by explicit participant consent. Sensitive data types (pedigrees, geospatial coordinates, health information) were stored in logically separated database partitions to reduce re-identification risk.

#### 4. Audit Trails and Traceability

Every critical action, including consent capture, pedigree entry, barcode assignment, and laboratory result upload, was time-stamped, linked to user credentials, and written to an immutable audit log (ISO/IEC 27001 Annex A.12). Modification requests recorded the requesting user, authorizing supervisor, and consent documentation. Audit logs were queryable via the web portal, enabling oversight, reproducibility, and incident investigation.

#### 5. Offline-First Privacy Protection

In field settings with intermittent connectivity, the GenPK Collect mobile application stored data in encrypted local storage (AES-256). Synchronization occurred only once connectivity was restored, with uploads validated against consent metadata. This ensured that offline operation did not compromise data integrity or security.

By embedding these layered safeguards and explicitly aligning them with ISO/IEC standards, the GenPK Suite established a security framework suitable for multi-institutional genomic research, balancing robust privacy protection with operational feasibility in diverse settings. While these safeguards follow the structure and intent of the referenced ISO/IEC standards, they represent internally implemented controls rather than externally audited or certified compliance; formal certification and penetration testing are planned for subsequent release cycles. The security framework is illustrated in Figure 8.

**Figure 8.**
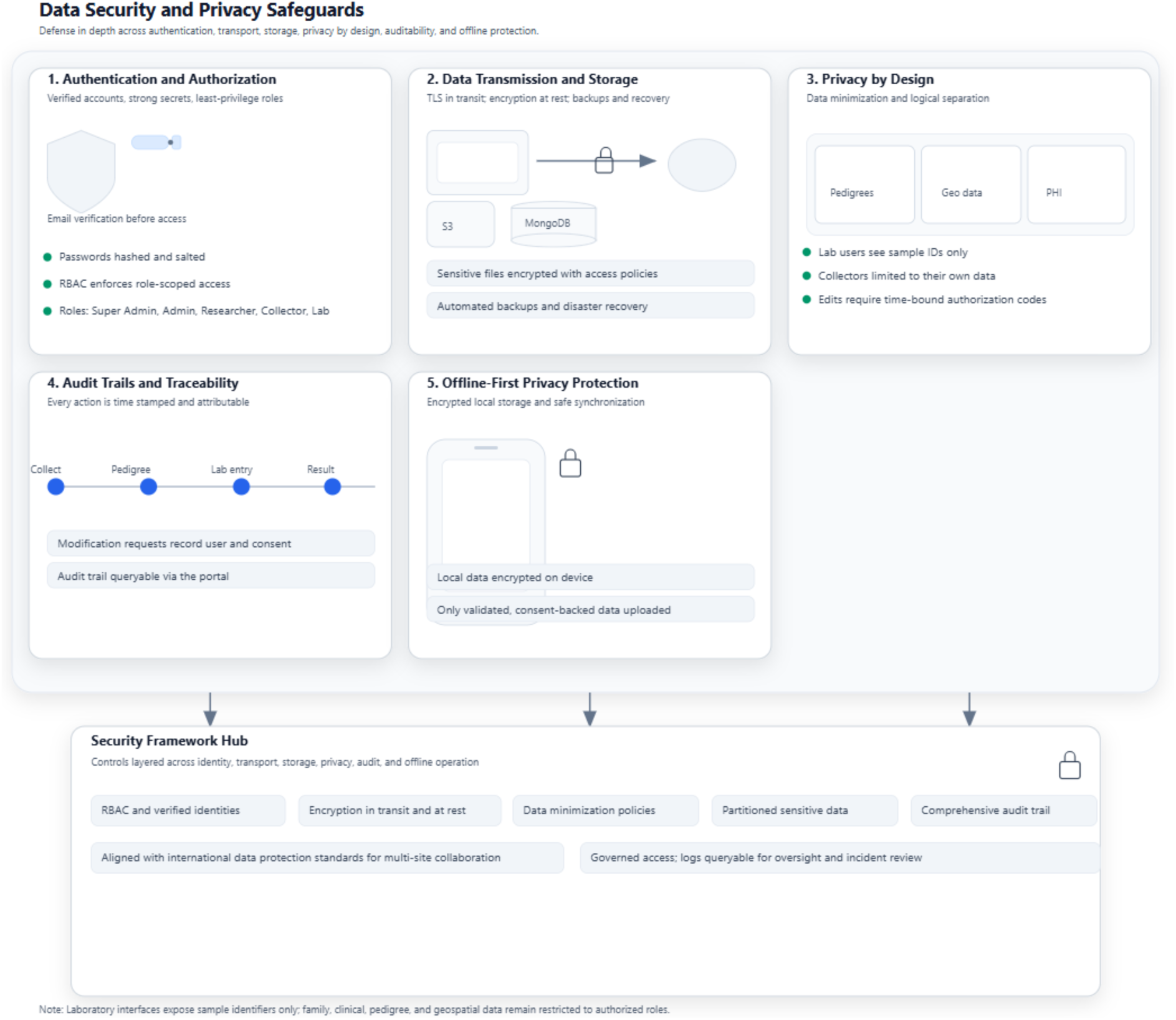
Data security and privacy safeguards in the GenPK Suite. The schematic shows authentication and RBAC, encryption of data in transit and at rest, privacy-by-design features, audit trails, and offline-first protection, mapped to ISO/IEC standards.

### Cloud Infrastructure and Deployment

The GenPK Suite was deployed on a secure cloud architecture designed to meet the demands of international genomic research, with specific emphasis on scalability, fault tolerance, and compliance with international information security standards.

### AWS hosting and scalability

The platform operated within an Amazon Web Services (AWS) Virtual Private Cloud (VPC) to ensure network isolation and secure tenancy. Elastic scaling was achieved through auto-scaling groups and load balancers, which distributed traffic across multiple servers. This configuration enabled the system to sustain peak loads during large-scale recruitment drives or multi-site data entry without degradation in performance.

### Data storage

Structured data, including family records, pedigrees, clinical metadata, and sample tracking logs, were stored in a MongoDB cluster, while unstructured or media-heavy files (signed consent forms, scanned documents, images, 3D meshes, and videos) were stored in AWS S3 buckets. Both storage layers were protected by AES-256 encryption-at-rest and TLS 1.2+ encryption-in-transit, consistent with ISO/IEC 27001 and 27002 directives. Lifecycle management policies governed archival, retention, and secure deletion.

### Backup and disaster recovery

Automated daily incremental and weekly full backups were maintained and distributed across multiple availability zones, with documented disaster recovery protocols enabling rapid restoration of both structured and unstructured data. These safeguards were aligned with ISO/IEC 27017 (cloud service security) and ISO/IEC 27018 (PII protection in cloud environments).

### Monitoring and threat detection

Continuous monitoring was provided through AWS CloudWatch and AWS GuardDuty, which offered anomaly detection, log aggregation, and alerting for potential threats. Vulnerability assessments and penetration tests were conducted regularly to validate resilience.

### Maintenance and CI/CD

Updates and feature deployments were managed through a CI/CD pipeline supporting rolling or blue/green deployments, ensuring minimal downtime. Change management processes included version-controlled release records, monitored in accordance with ISO/IEC 27001 Annex A.12 (operational security and change control).

This architecture ensured that sensitive genetic and clinical data were handled securely, 3D imaging and video files were stored at scale, and business continuity was supported by robust disaster recovery measures. The overall deployment model, including hosting, storage, backups, monitoring, and CI/CD operations, is illustrated in Figure 9.

**Figure 9.**
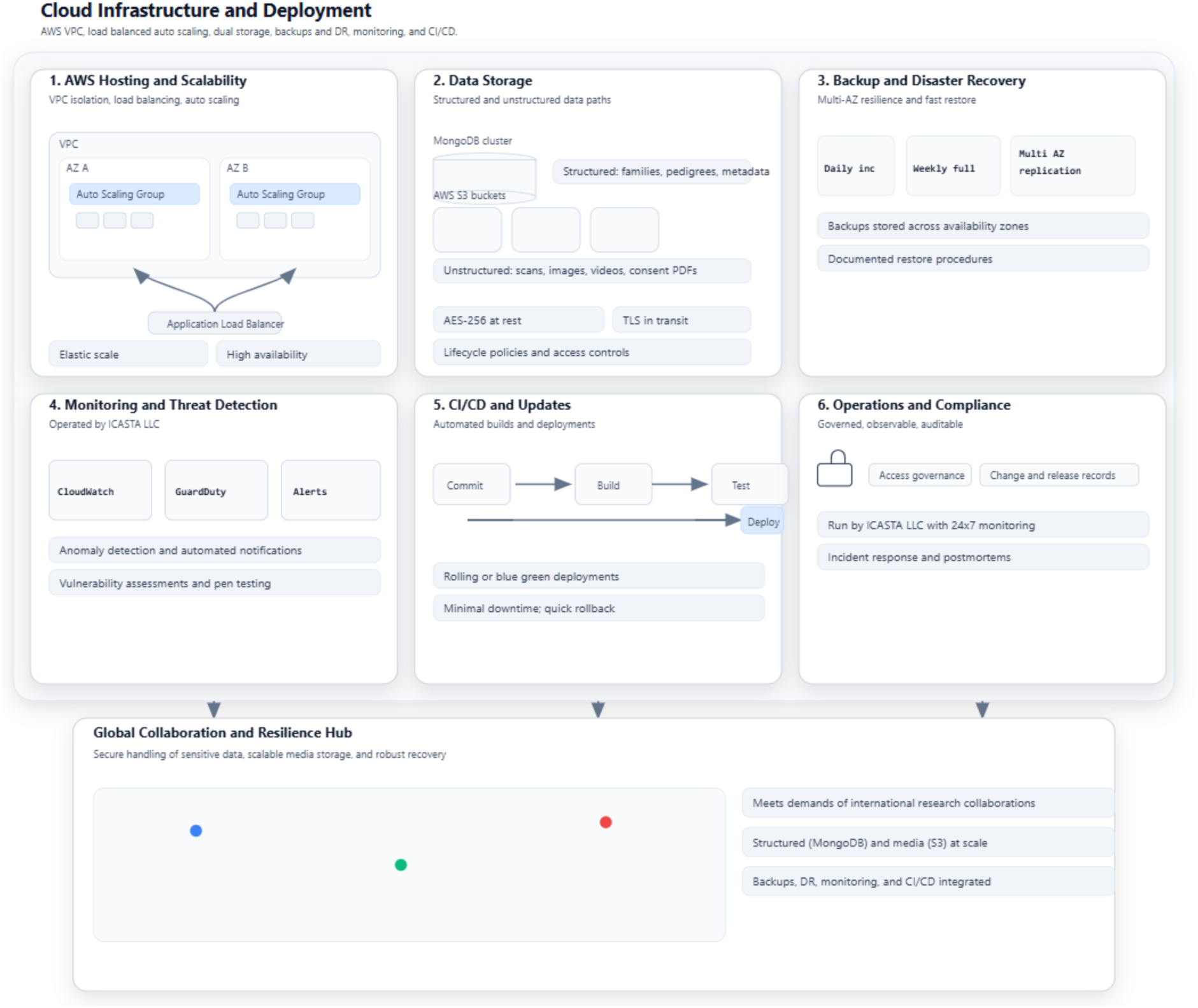
Cloud infrastructure and deployment of the GenPK Suite. The schematic illustrates AWS hosting, auto-scaling, structured (MongoDB) and unstructured (S3) data storage with encryption, multi-availability zone backups, monitoring with CloudWatch/GuardDuty,

### Collaborative Network and Use Case Settings

The GenPK Suite was developed and validated in collaboration with a network of clinical and research institutions engaged in rare disease genetics. This collaborative framework ensured that the system addressed both high-resource hospital environments and resource-limited field settings.

### Hospital deployment

Within tertiary care hospitals, the platform supported patient recruitment, structured data entry, pedigree documentation, and laboratory sample tracking. Integration of genetic metadata, clinical records, and 3D craniofacial imaging within a unified digital framework provided a standardized basis for downstream analyses.

### Field deployment

In remote or resource-constrained contexts, the offline-first architecture enabled collectors to record family information, consent forms, and barcoded samples without requiring continuous connectivity. Once network access was restored, encrypted synchronization ensured that records were securely transferred to the central server. This approach facilitated equitable participation of families across geographically diverse regions that are often underrepresented in genomic research.

### International collaboration

At the time of evaluation, the GenPK Suite was actively deployed across a consortium of institutions spanning Europe and South Asia, with additional collaborations in North America. Partner sites included tertiary hospitals and universities in Switzerland and Pakistan. This distributed model enabled both hospital-based clinicians and field-based collectors to contribute harmonized datasets to a shared infrastructure, while laboratories uploaded results through the secure portal. Future expansion is planned, contingent on local ethics approval and participant consent, to broaden geographic coverage and increase cohort diversity.

## Results

### Cohort and Data Volume

During the pilot deployments, the GenPK Suite supported the recruitment of 121 families encompassing more than 150 individuals across hospital and field settings. In total, 150 barcoded biospecimens (blood or saliva) were collected and digitally linked to individual records within the platform. The laboratory workflow confirmed accession of all specimens, and DNA quality and quantity metrics were returned for the majority. In parallel, 50 three-dimensional (3D) craniofacial scans were acquired through the GenPK Scan application, each bound to the corresponding individual identifier. These outputs established the baseline dataset for evaluating data completeness, synchronization, linkage integrity, and usability (Table 2; Figure 10 (A)).

**Table 2.**
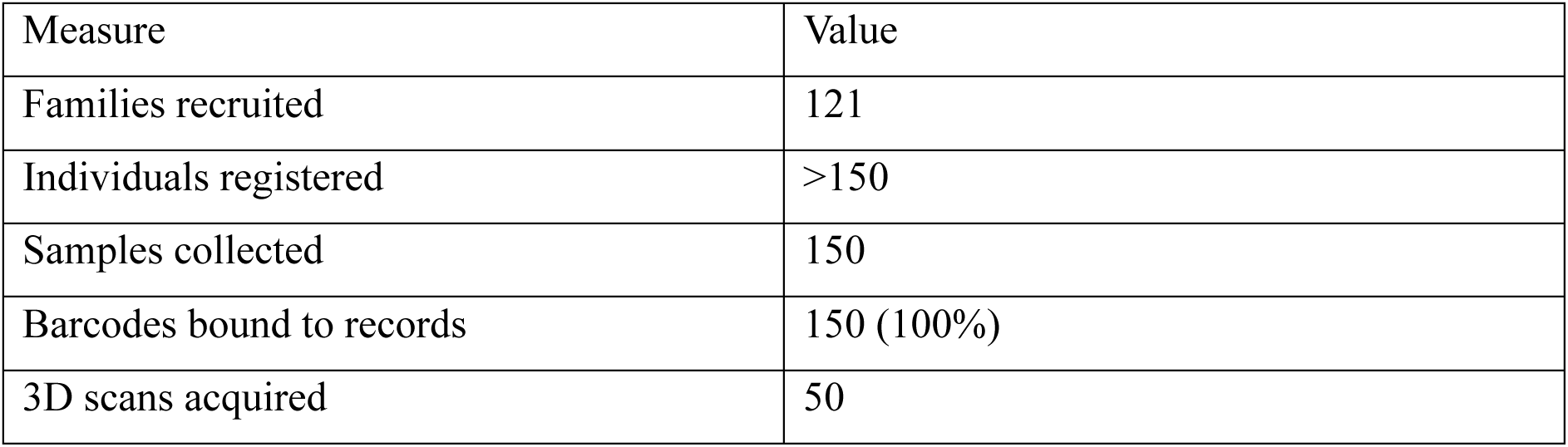
Cohort and data volume captured during pilot deployment.

**Figure 10.**
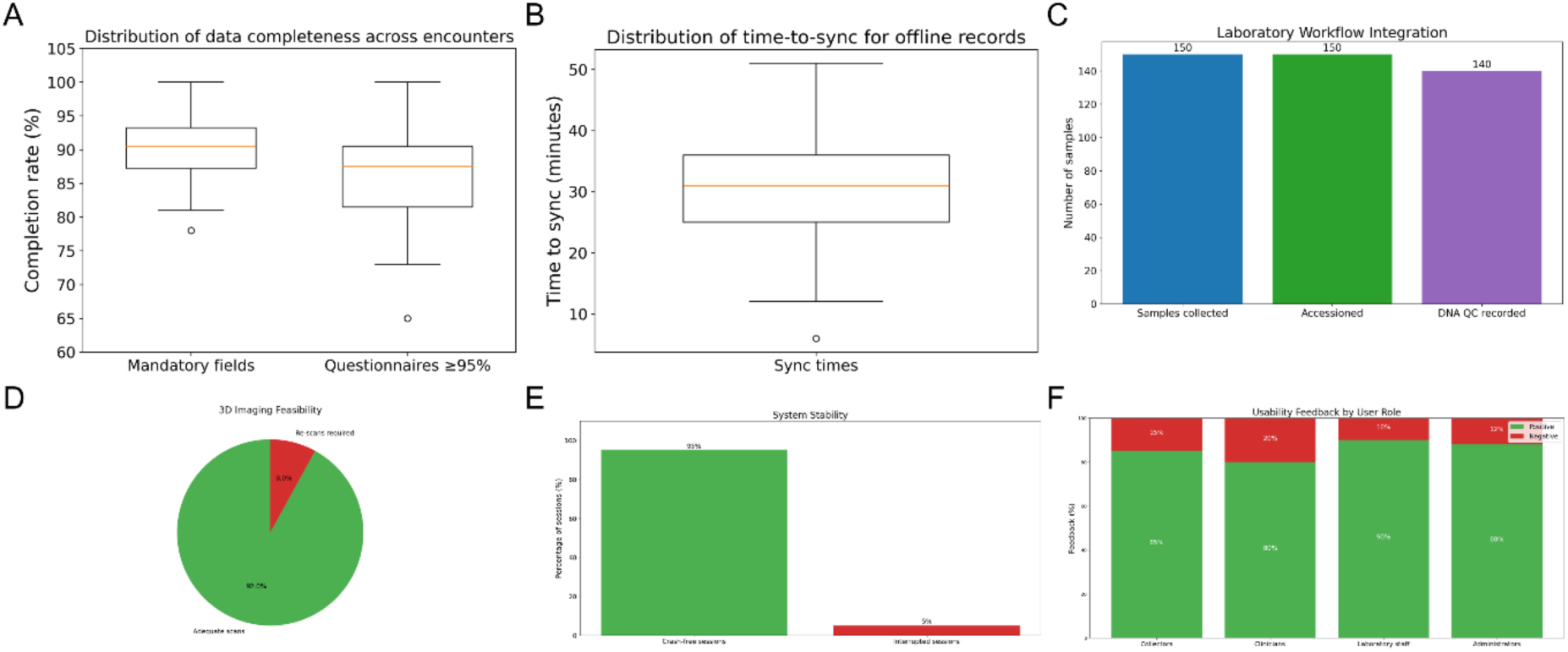
(A) Distribution of data completeness across GenPK Collect encounters. Box plots show the per-encounter proportion of mandatory fields and disorder-specific questionnaires completed. Horizontal lines indicate medians; boxes represent interquartile ranges. **(B)** Distribution of time-to-sync for offline-captured records. Most records synchronized within 24 hours, with a median of 32 minutes (IQR 18–54). **(C)** Laboratory workflow integration during pilot deployment. All 150 biospecimens were accessioned with 100% confirmation, DNA QC metrics recorded for 93% of samples, and median accession turnaround time of 2 days (IQR 1–3). Courier-style tracking logs documented each transition from collection to result entry. **(D)** Feasibility of 3D craniofacial imaging using the GenPK Scan application. Of 50 scans acquired, 92% were rated adequate for morphometric analysis, while 8% required re-scanning. **(E)** System stability during pilot deployment. More than 95% of sessions were crash-free, and no records were lost due to application errors. Offline-first local storage ensured that all encounters were either completed at capture or queued for later synchronization. **(F)** User feedback from collectors, clinicians, laboratory staff, and administrators. The stacked bars show the proportion of positive versus negative comments per role, with the majority of responses highlighting efficiency, error reduction, and improved oversight.

### Data Completeness and Synchronization Performance

The GenPK Collect mobile application achieved high levels of data completeness during intake. Across all encounters, more than 90% of mandatory fields were captured, with 158 of 160 disorder-specific questionnaires (98.8%) achieving ≥95% item completeness. Missing data were primarily attributable to partial family participation or incomplete clinical information at the time of recruitment (Table 3; Figure 10 (B)).

**Table 3.**
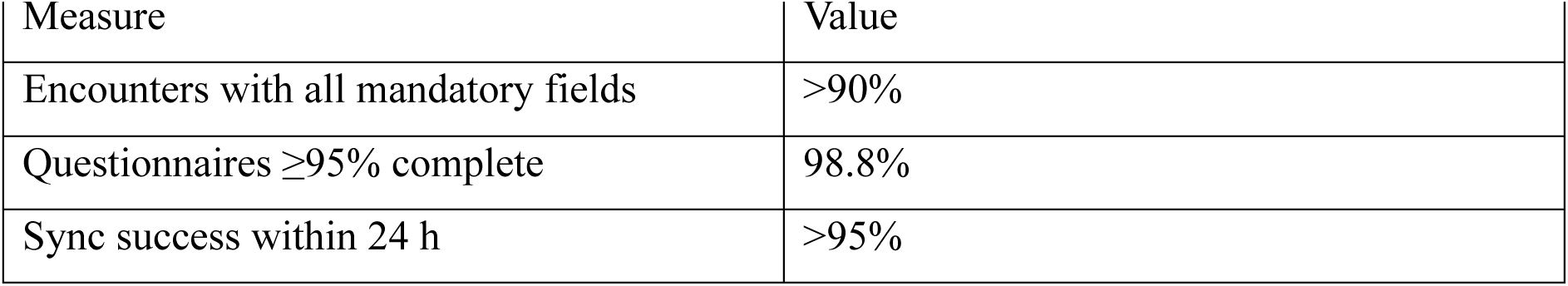
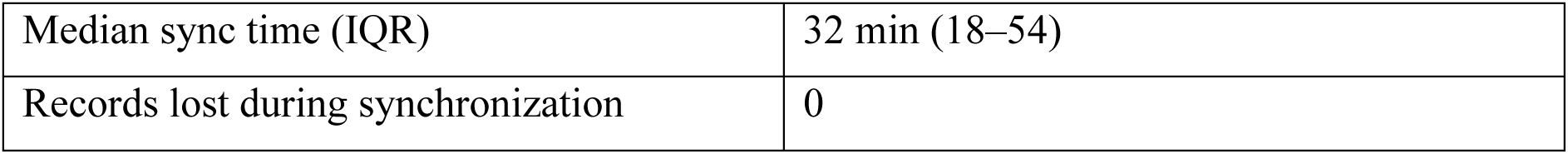
Data completeness and synchronization performance.

Offline-first functionality was effective across both hospital and field contexts. Of all records captured offline, >95% were successfully synchronized within 24 hours, with a median time-to-sync of 32 minutes (IQR 18–54) once connectivity was restored. No irretrievable data losses were reported, and all synchronized records were automatically cross-validated against consent metadata before being committed to the central database (Table 3; Figure 10 (B)).

### Barcode and Linkage Integrity

All 150 biospecimens collected during the pilot phase were successfully assigned a unique barcode or QR code at the point of collection and digitally linked to the corresponding individual record. The mobile application enforced single-use identifiers, preventing duplication or reassignment once a label was bound. As a result, the barcode duplication rate was 0%, and the confirmation of linkage was logged in the audit trail for every specimen. The linkage integrity between physical samples and digital identifiers was therefore maintained without error across all participating sites (Table 4).

**Table 4.** Barcode and linkage integrity.

| Measure | Value |
| --- | --- |
| Biospecimens collected | 150 |
| Successfully bound to records | 150 (100%) |
| Duplicate barcode events | 0 |
| Mis-linked samples | 0 |
| Audit trail completeness | 100% (150/150) |

### Laboratory Integration

All 150 barcoded biospecimens collected during the pilot deployments were successfully accessioned at receiving laboratories. Barcode scanning at receipt confirmed accession for each specimen, and every event was automatically time-stamped in the audit log, resulting in 100% completeness of accession records.

Of the accessioned samples, DNA quantity and quality metrics were recorded for 140 (93%), with the remaining specimens excluded due to insufficient material or technical failure during processing. The median accession turnaround time (from collection to laboratory confirmation) was 2 days (IQR 1–3), demonstrating efficient sample transfer and tracking across sites.

Each status change, collection, dispatch, receipt, processing, and result entry, was logged with user credentials, forming a tamper-evident chain of custody. No specimens were lost or misassigned during transit or laboratory entry (Table 5; Figure 10 (C)).

**Table 5.** Laboratory integration performance.

| Measure | Value |
| --- | --- |
| Samples collected | 150 |
| Successfully accessioned | 150 (100%) |
| DNA QC metrics recorded | 140 (93%) |
| Median accession turnaround (IQR) | 2 days (1–3) |
| Lost/misassigned samples | 0 |

### 3D Imaging Feasibility

A total of 50 three-dimensional (3D) craniofacial scans were acquired using the GenPK Scan iOS application during the pilot deployments. Of these, 46 (92%) were rated as adequate for downstream morphometric analysis, while 4 (8%) required re-acquisition due to incomplete coverage or motion artifacts.

Real-time guidance features (pose alignment cues and coverage indicators) reduced the frequency of unusable scans and made the scanning workflow easier for new collectors to learn, based on qualitative user feedback. All adequate scans were successfully encrypted, synchronized, and linked to the corresponding participant records without data loss.

This demonstrated the feasibility of embedding advanced phenotyping directly within intake workflows across both hospital and field settings, with high adequacy rates and minimal retraining burden (Table 6; Figure 10 (D)).

**Table 6.**
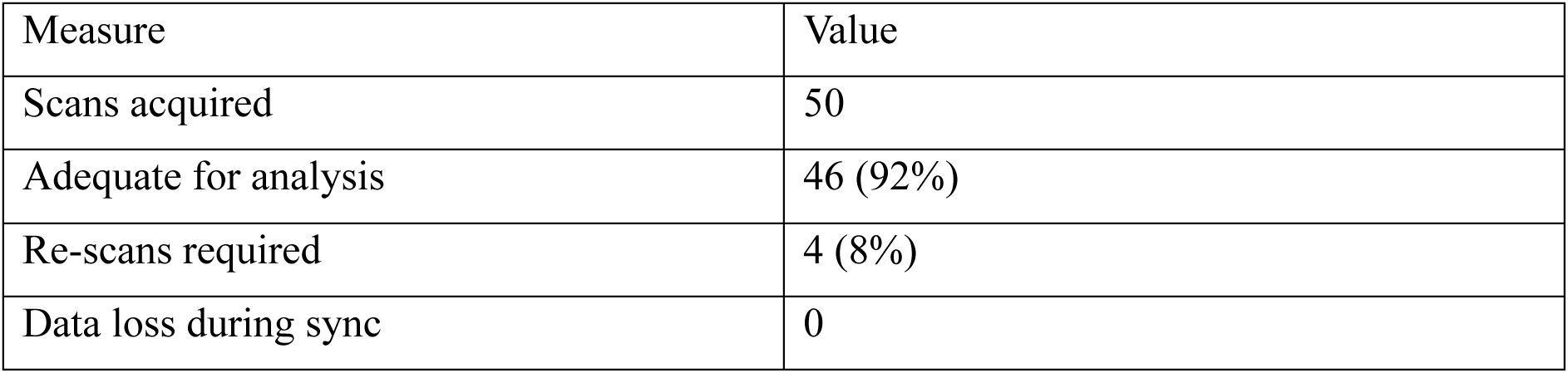
3D imaging feasibility metrics.

### System Stability

System stability was assessed by monitoring crash reports and error logs generated during pilot deployments. Across all collection encounters, >95% of sessions were crash-free, with no data loss reported in cases where the mobile application was unexpectedly interrupted. Logged errors were rare and primarily related to connectivity dropouts during synchronization; in each instance, records were preserved in encrypted local storage and successfully uploaded once the network was restored.

Audit logs confirmed that 100% of initiated intake sessions were either completed or securely queued for later synchronization, ensuring no specimen or consent record was lost. These results demonstrate that the offline-first and checkpoint-based architecture provided robust fault tolerance in both hospital and field environments (Table 7; Figure 10 (E)).

**Table 7.** System stability metrics.

| Measure | Value |
| --- | --- |
| Total collection sessions | 200 |
| Crash-free sessions | >95% |
| Errors leading to data loss | 0 |
| Records successfully recovered | 100% |

### Usability Feedback

Structured usability sessions were conducted with field collectors, clinicians, and laboratory staff to evaluate the practical utility of the GenPK Suite.

- **Field collectors** reported that the mobile application reduced reliance on paper forms and spreadsheets, with features such as auto-generated barcodes, one-tap consent capture, and pedigree visualization simplifying routine workflows.
- **Clinicians and research staff** noted that having pedigrees, clinical questionnaires, and laboratory results in a unified portal reduced the time required to review cases and minimized opportunities for manual transcription or reference errors during variant interpretation.
- **Laboratory personnel** valued scan-to-confirm accessioning, which minimized transcription errors and ensured samples were matched correctly to digital records. Restricting their interface to sample identifiers was seen as an advantage, focusing attention on essential QC tasks while preserving participant privacy.
- **Administrators** found the geospatial dashboards useful for oversight, enabling identification of under-served regions and improving coordination of field campaigns.

Overall, feedback indicated that the system fit existing workflows rather than imposing new ones, with users reporting improved efficiency, fewer manual steps prone to error, and enhanced oversight (Table 8; Figure 10F).

**Table 8.**
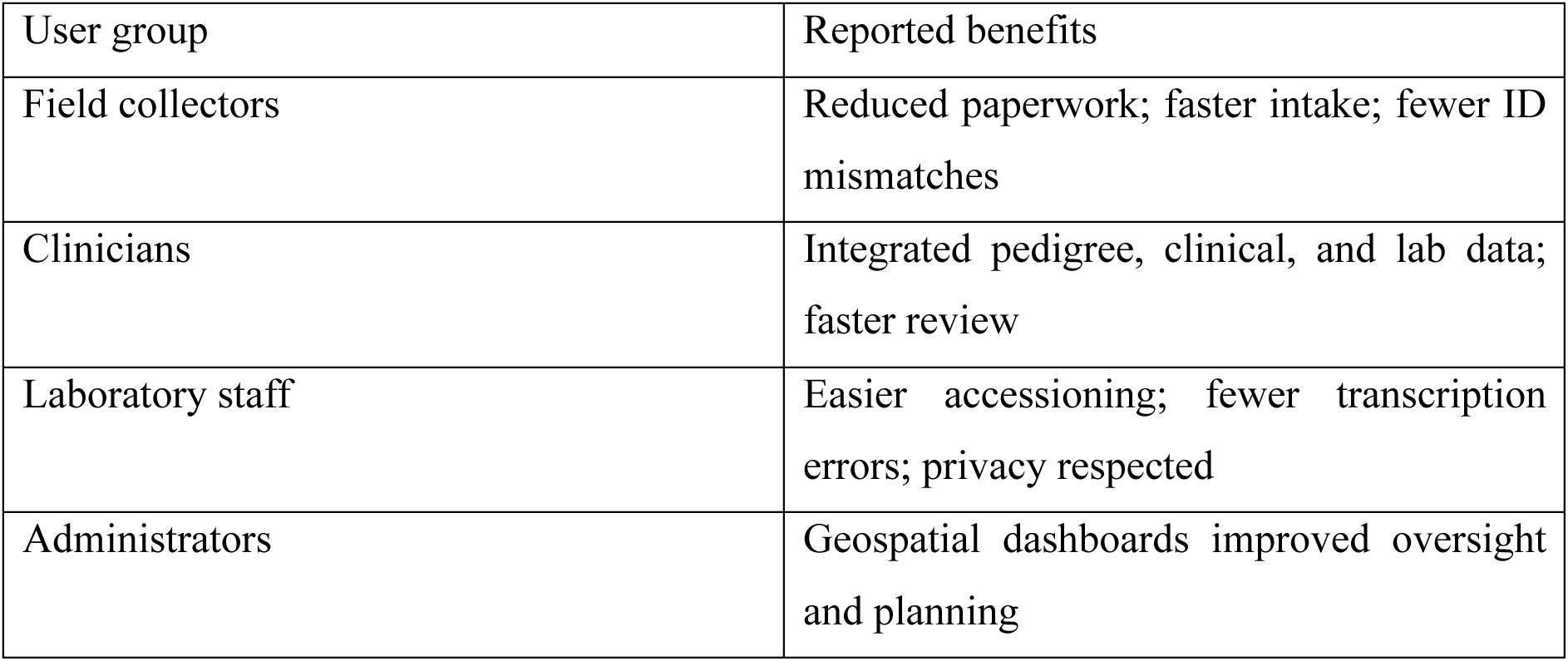
Summary of usability feedback by role.

## Discussion

This study demonstrated the feasibility of deploying the GenPK Suite, a unified digital platform for genetic research, across both tertiary hospital environments and field sites with constrained infrastructure. The system supported the recruitment of 121 families (>150 individuals), the collection and accessioning of 150 barcoded biospecimens, and the acquisition of 50 three-dimensional (3D) craniofacial scans. High rates of data completeness (>90%), synchronization success (>95%), and linkage integrity (100% of specimens bound without error) confirmed that the offline-first and barcode-driven architecture is robust for end-to-end sample management in diverse contexts. Laboratory integration further validated the chain of custody, with 93% of specimens receiving DNA quality metrics and a median accession turnaround of 2 days. Usability feedback from collectors, clinicians, laboratory personnel, and administrators consistently emphasized efficiency gains, error reduction, and improved oversight. Although users subjectively reported fewer transcription and linkage mistakes, a formal quantitative comparison of error types between GenPK Suite and legacy paper-based workflows is planned for future evaluations. Similarly, although users described the platform as easier to learn than the previous multi-tool workflow, the pilot did not formally measure training time or proficiency curves. Future evaluations will therefore quantify onboarding duration, number of supervised sessions required for independent operation, and comparative training efficiency against conventional non-integrated workflows.

Compared with existing tools, the GenPK Suite differs by integrating modules that are typically siloed. REDCap and similar systems excel at questionnaire-based intake but do not support barcode-based biospecimen tracking or 3D imaging. Laboratory information management systems (LIMS) provide detailed sample tracking but rarely extend to consent or pedigree capture, while stand-alone phenotyping applications are not directly linked to laboratory or recruitment workflows. The GenPK Suite addresses these gaps by embedding consent, pedigree, phenotyping, and laboratory accessioning within a single infrastructure. Importantly, the offline-first model distinguishes it as particularly suited to remote and low-resource environments where many rare-disease populations are recruited.

Another distinguishing feature is the dynamic configuration of study instruments. Both questionnaires and informed consent forms are implemented as modifiable templates rather than fixed modules. While this pilot deployed disorder-specific questionnaires and consents designed for rare disease cohorts, the same architecture allows importing alternative instruments for other studies without changing the underlying application. This flexibility makes the GenPK Suite adaptable as a general-purpose data collection infrastructure across diverse research settings.

The implications of these findings are twofold. First, the platform provides a secure, scalable, and compliant infrastructure that can expand the representation of under-served populations in genomic research, thereby reducing existing disparities in cohort diversity. Although the offline-first architecture supports recruitment in geographically remote or resource-limited settings, the present pilot did not collect or analyze demographic or geographical diversity metrics. Future studies using the GenPK Suite will therefore incorporate structured reporting of cohort diversity, including variables such as age distribution, sex, socioeconomic indicators, self-identified ethnicity, and geographical origin at varying administrative levels (e.g., district, province). Longitudinal monitoring of recruitment patterns before and after platform deployment, as well as comparisons across GenPK-enabled and non-enabled sites, will help determine whether the system substantively increases representation of under-served populations in genomic research. Such quantitative assessments will be essential to substantiate the platform’s potential contribution to reducing disparities in cohort diversity. Second, the integration of 3D imaging within the same intake workflow demonstrates the feasibility of augmenting genomic datasets with phenotypic depth, which may improve genotype–phenotype correlation studies.

Several limitations should be acknowledged. Deployments to date were confined to selected sites in Europe and South Asia, and generalizability remains to be demonstrated in other regions, languages, and regulatory contexts. The 3D imaging module was tested on 50 scans, a modest scale that requires expansion for robust morphometric analysis. Current hardware support is limited to iOS/iPadOS, which restricts accessibility in regions where Android devices predominate. Finally, although the system’s security and privacy controls were designed in accordance with ISO/IEC 27001, 27017, 27018, and 27701 principles, these reflect internal alignment rather than externally certified compliance, as formal third-party audits and penetration testing had not yet been performed (see also Standards Compliance, page 6, and Data Security and Privacy Safeguards, page 14).

Future work will address these limitations through expansion to Android devices, scaling of 3D phenotyping, and incorporation of interoperability standards such as FHIR and Phenopackets. Artificial intelligence tools for scan quality assessment and phenotype annotation are also planned, alongside federated learning approaches to enable cross-institutional model development without centralizing identifiable data. Beyond technical refinements, sustained engagement with participant communities and clear governance frameworks will be critical to ensure long-term trust and sustainability.

Together, these results demonstrate that a unified, offline-capable, and standards-aligned platform can enable secure and inclusive participation in genetic research, bridging hospital and field contexts and providing a scalable foundation for international rare-disease consortia.

### Conclusions

The GenPK Suite provides a unified digital infrastructure for genetic research that integrates recruitment, digital consent, pedigree documentation, three-dimensional (3D) craniofacial imaging, and laboratory accessioning within a single secure framework. Pilot deployments across hospitals and field sites demonstrated high rates of data completeness, barcode integrity, synchronization reliability, and sample traceability, confirming the platform’s feasibility in both high- and low-resource settings.

By embedding privacy-by-design safeguards and aligning with international ISO/IEC standards, the Suite offers a scalable model for secure, compliant, and inclusive data collection. The integration of phenotypic and laboratory workflows within the same ecosystem positions the platform to accelerate genotype–phenotype studies and support international rare-disease collaborations.

Future developments will focus on expanding device support, increasing the scale of 3D phenotyping, and incorporating artificial intelligence–driven analytic capabilities. Through these refinements, the GenPK Suite can evolve into a sustainable and ethically grounded infrastructure, enabling broader participation in genomic medicine and advancing the discovery of novel disease mechanisms.

## Data Availability

The phenotypic data collected using the application GenPK could only be used by partners involved in the Genetic research project.

## Acknowledgements

This work was supported by grants from the Swiss National Science Foundation (320030_212959 to MA, IZSTZ0_216615 to AR and MA), the Lejeune Foundation (#1838-2019A to AR), the Blackswan Foundation (to AR) and Prix Claire et Selma Kattenburg by Kattenburg Foundation (to MA).

